# Childhood listening and associated cognitive difficulties persist into adolescence

**DOI:** 10.1101/2022.08.11.22278673

**Authors:** Katsuaki Kojima, Li Lin, Lauren Petley, Nathan Clevenger, Audrey Perdew, Mark Bodik, Chelsea M. Blankenship, Lina Motlagh Zadeh, Lisa L. Hunter, David R. Moore

## Abstract

**Objective:** Listening difficulty (LiD) refers to the challenges individuals face when trying to hear and comprehend speech and other sounds. LiD can arise from various sources, such as hearing sensitivity, language comprehension, cognitive function, or auditory processing. Although some children with LiD have hearing loss, many have clinically normal audiometric thresholds. To determine the impact of hearing and cognitive factors on LiD in children with a clinically normal audiogram, we conducted a longitudinal study. The Evaluation of Children’s Listening & Processing Skills (ECLiPS), a validated and standardized caregiver evaluation tool, was used to group participants as either LiD or typically developing (TD). Our previous study aimed to characterize LiD in 6- to 13-year-old children during the project’s baseline, cross-sectional phase. We found that children with LiD needed a higher signal-to-noise ratio during speech-in-speech tests and scored lower on all assessed components of the NIH Cognition Toolbox than TD children. The primary goal of this study was to examine if the differences between LiD and TD groups are temporary or enduring throughout childhood.

**Design:** This longitudinal study had three data collection waves for children with LiD and TD aged 6-13 years at Wave 1, followed by assessments at 2-year (Wave 2) and 4-year (Wave 3) intervals. Primary analysis focused on data from Waves 1 and 2. Secondary analysis encompassed all three waves despite high attrition at Wave 3. Caregivers completed the ECLiPS, while participants completed the Listening in Spatialized Noise Sentences test (LiSN-S) and the NIH Toolbox Cognition Battery during each wave. The analysis consisted of 1) examining longitudinal differences between TD and LiD groups in demographics, listening, auditory, and cognitive function; 2) identifying functional domains contributing to LiD; and 3) test-retest reliability of measures across waves. Mixed-effect models were employed to analyze longitudinal data.

**Results:** The study enrolled 169 participants, with 147, 100, and 31 children completing the required testing during Waves 1, 2, and 3, respectively. The mean ages at these waves were 9.5, 12.0, and 14.0 years. On average, children with LiD consistently underperformed TD children in auditory and cognitive tasks across all waves. Maternal education, auditory and, especially, cognitive abilities independently predicted caregiver-reported listening skills. Significant correlations between Waves 1 and 2 confirmed high, long-term reliability. Secondary analysis of Wave 3 was consistent with the primary analyses of Waves 1 and 2, reinforcing the enduring nature of listening difficulties.

**Conclusion:** Children with LiD and clinically normal audiograms experience persistent auditory, listening, and cognitive challenges through at least adolescence. The degree of LiD can be independently predicted by maternal education, cognitive processing, and spatial listening skills. This study underscores the importance of early detection and intervention for childhood LiD and, for the first time, highlights the role of socioeconomic factors as contributors to these challenges.

## INTRODUCTION

Listening is commonly defined as an active form of hearing (Sweetow and Henderson-Sabes 2004). It facilitates an accurate understanding of speech, a key component of everyday communication. Successful communication allows healthy language development, as seen in improved language outcomes in children with hearing loss who received early interventions (Pimperton and Kennedy 2012; Yoshinaga-Itano et al. 1998; Moeller 2000; Dettman et al. 2007). Problems with listening may similarly predispose children to challenges in language acquisition. It is difficult, however, to determine the direction of the causal pathway because children with primary language or other cognitive problems may have difficulties comprehending speech that prevents them from active hearing (listening). Listening difficulty (LiD) describes difficulty hearing and understanding speech and other suprathreshold sounds (Petley et al. 2021; Dillon and Cameron 2021; Sharma et al. 2014). LiD is an umbrella term that describes symptoms; the underlying causes of LiD may be problems in multiple domains, including hearing, language, cognition, and auditory processing (Dillon and Cameron 2021; Sharma et al. 2009; Tomlin et al. 2015). Although some children presenting with LiD have hearing impairments, as indicated by elevated audiometric thresholds, other children with LiD have clinically normal audiometric sensitivity (Hind et al. 2011). To clarify the contribution of hearing and cognitive factors to LiD for individuals with clinically normal audiograms, we prospectively studied children with LiD, identified using a validated and standardized caregiver questionnaire, Evaluation of Children’s Listening & Processing Skills (ECLiPS) (Barry and Moore 2021).

In the Sensitive Indicators of Childhood Listening Difficulties (SICLiD) study, we characterized LiD in a cohort of 6- to 13-year-old children during a baseline, cross-sectional phase of the project, using caregiver questionnaires and a battery of audiological, auditory processing, and cognitive tests (Petley et al. 2021; Hunter et al. 2021). We found no group differences in audiological measures (pure-tone audiometry, tympanometry, and middle ear muscle reflexes) between LiD and typically developing (TD) children (Hunter et al. 2021). However, children with LiD required a higher signal-to-noise ratio during speech hearing in noise testing (Listening in Spatialized Noise-Sentence, LiSN-S (Cameron and Dillon 2007)), had abnormal auditory processing measures tested using SCAN-3:C (Keith 2009), and lower cognitive scores in all tested components of the NIH Cognition Toolbox (Weintraub et al. 2013b) relative to their TD peers (Petley et al. 2021).

Whether LiD persists from childhood into adulthood or resolves over time remains unclear. Answering this question is essential for informing clinical management and understanding the mechanisms and consequences of LiD. If LiD is a persistent problem, more comprehensive assessment and aggressive treatment may be warranted compared to a transient problem of childhood. Some evidence suggests LiD may be persistent - young adults with a history of LiD in childhood reported ongoing communication challenges nearly 10 years after initial diagnosis (Del Zoppo et al. 2015). However, quantitative longitudinal data tracking changes in listening, auditory, and cognitive functioning in children diagnosed with LiD earlier in life is lacking. As part of the SICLiD study, we performed behavioral testing in the same cohort of children every two years to assess longitudinal changes.

Socioeconomic status has been associated with childhood cognitive performance (Christensen et al. 2014), language development (Pungello et al. 2009), mental health problems (Reiss 2013), and health disparities (Fiscella and Williams 2004). However, the impact of social factors on LiD remains unclear. Given the association between LiD and cognitive performance, we hypothesized that the degree of LiD may also be associated with social factors. In a prior cross-sectional study, we developed a model showing that SCAN-3:C, LiSN-S, and NIH Cognition Toolbox scores partially accounted for caregiver reports of LiD (Petley et al. 2021). While social factors were evaluated, they were not incorporated into the final model. Longitudinal data allowed us to assess social and other demographic factors across time and with increased statistical power.

Evaluating longitudinal cognitive performance during development can be challenging because of the different rates of development and potential practice effects (Sullivan et al. 2017). In the SICLiD study, we used the NIH Cognition Toolbox to assess cognition. Weintraub et al. (2013b) showed that the Toolbox test results had high reliability in pediatric samples with short testing intervals between 7 and 21 days, with intraclass correlations (ICCs) between 0.76 and 0.99. However, recent literature on the reliability for longer intervals (> 1 year) has been mixed, with ICCs between 0.24 and 0.85 (Taylor et al. 2022) and 0.43 and 0.82 (Anokhin et al. 2022). The SICLiD study enabled evaluation of the reliability of the NIH Cognition Toolbox over the years.

We had three aims in the current study. The primary aim was to evaluate longitudinal changes in listening, auditory, and cognitive functions of children with LiD relative to control children. We hypothesized that LiD would be long-lasting because LiD was seen across ages (between 6 and 13 years old) in our Wave 1, cross-sectional data (Petley et al., 2021), and because of the general persistence through adolescence and into adulthood of developmental problems associated with LiD, such as attention (Hechtman et al. 2016; Roy et al. 2017), reading (Shaywitz et al. 1999; Lohvansuu et al. 2021), and language (St Clair et al. 2011). Second, building on the previous model (Petley et al., 2021), we aimed to develop an enhanced model of contributors to LiD using longitudinal data. Finally, we examined long-term test-retest reliability of the NIH Cognition Toolbox, ECLiPS, and LiSN-S by using prospectively collected data from children with and without LiD.

## MATERIALS AND METHODS

The Institutional Review Board of Cincinnati Children’s Hospital (CCH) Research Foundation approved this study.

### Participants

LiD and TD children were 6 to 13 years old at enrollment. The LiD group was recruited from children referred to CCH for audiological evaluation and from the community. All the participants in the TD group were recruited from the community. Recruitment from the community involved flyers posted in relevant CCH clinics (Audiology, Pediatrics, Speech-Language Pathology) and emails sent to all CCH employees and families interested in research. Interested caregivers completed (i) a background questionnaire and (ii) the ECLiPS (Barry and Moore 2021) to confirm eligibility for the LiD or TD group. Total standardized ECLiPS scores of ≥ 7 defined the TD group, and scores < 7 defined the LiD group. However, four children with LiD had a previous audiologic diagnosis of auditory processing disorder (APD) but scored 7 - 9 on the ECLiPS at enrollment. APD is a hypothesized source of LiD (Dillon and Cameron 2021). The inclusion criterion for both groups was English as the child’s native language. Exclusion criteria for both groups were reported neurologic (e.g., epilepsy or stroke), psychiatric (e.g., attention deficits hyperactivity disorder (ADHD) requiring medical treatment), or intellectual conditions that would prevent or restrict their ability to complete testing procedures. The TD group additionally could not have any reported listening difficulty, or cognitive or learning disorder (e.g., developmental delay, attention, language, reading, or high-functioning autistic diagnoses). Eligibility was determined based on caregiver responses to the background questionnaire. Groups were age-matched by proportional sampling (Table 1).

**Table 1.** Participant numbers and demographic information.

|  | Wave 1 |  | Wave 2 |  | Wave 3 |  |
| --- | --- | --- | --- | --- | --- | --- |
|  | TD | LiD | TD | LiD | TD | LiD |
| N | 79 | 68 | 48 | 52 | 15 | 16 |
| Age, year (SD) | 9.3 (2.0) | 9.7 (1.9) | 12.1 (2.2) | 11.9 (2.0) | 13.4 (1.9) | 14.5 (2.2) |
| Female, n (%) | 33 (42) | 24 (35) | 24 (50) | 16 (31) | 6 (40) | 6 (38) |
| Maternal education, n (%) |  |  |  |  |  |  |
| High school | 1 (1) | 12 (18) | 1 (2) | 9 (18.4) | 0 (0) | 3 (19) |
| College | 8 (10) | 17 (25) | 3 (6) | 15 (30.6) | 0 (0) | 6 (38) |
| Bachelor | 50 (63) | 25 (37) | 31 (65) | 16 (32.7) | 11 (73) | 3 (19) |
| Post-grad | 20 (25) | 14 (21) | 13 (27) | 9 (18.4) | 4 (27) | 4 (25) |
| Race, non-white (%) | 13 (16) | 17 (25) | 5 (10) | 16 (31) | 1 (7) | 9 (56) |
| Ethnicity, Hispanic or Latino (%) | 3 (4) | 4 (6) | 2 (4) | 3 (6) | 1 (7) | 2 (12) |

One hundred sixty-six participants (74 with LiD, 92 TD) were enrolled; 19 withdrew or otherwise exited the study (6 with LiD, 13 TD). Reasons for withdrawal were schedule conflicts, transportation availability, and becoming ineligible after enrollment. The remaining 147 participants (68 with LiD, 79 TD) completed the assessments at baseline (Wave 1). After the initial study visit, we contacted the participants after ∼2 years (Wave 2) and 4 years (Wave 3) to collect longitudinal data. A total of 100 (52 with LiD, 48 TD) and 31 (16 with LiD, 15 TD) children participated in Wave 2 and 3, respectively (Table 1). The sample size for Wave 3 was notably smaller than other waves, partly because of the COVID-19 pandemic. All participants had clinically normal hearing bilaterally, as detailed below.

### Procedure

All caregivers of eligible and interested participants reviewed the informed consent form with a study staff member and discussed the purpose, procedures, risks and benefits, duration, and expectations involved in the study. According to institutional policy, children aged 11 and above also assented using a child-friendly version of the consent document. All participants received financial compensation for their participation. Each participant completed audiometric and behavioral testing during the study visit, as listed below. Several additional tests, including brainstem and cortical evoked responses and structural and functional MRI, were also performed during the visit as part of the SICLiD study. The results of these additional tests are beyond the scope and not included in the current manuscript. Some results of these additional tests have been reported elsewhere (Hunter et al. 2021; Stewart et al. 2022; Hunter et al. 2023).

### Background Questionnaire

Study staff developed a background questionnaire to collect information about the participants, including demographics, medical and surgical history, and socioeconomic status. This questionnaire lacks reliability or validity data because of its basic nature. Specific information collected by this questionnaire was age, sex, race, ethnicity, maternal education (a proxy for socioeconomic status (Sherar et al. 2016)), primary language used at home, and diagnosis and treatment received for otologic, neurologic, behavioral, developmental, and psychiatric problems. The questionnaire included specific developmental conditions as examples, such as ADHD, autism spectrum disorder, and dyslexia. Caregivers completed the same background questionnaire at each Wave. Demographic variables at each time point are summarized in Table 1.

### Caregiver Evaluation of Children’s Listening

The ECLiPS (Barry and Moore 2021) was administered at each wave to assess participants’ listening and communication abilities. This questionnaire consists of 38 straightforward statements (items) describing behaviors commonly observed in children. Caregivers rated their child on a five-point scale, ranging from strongly disagree to strongly agree, based on their agreement with each statement. These ratings were averaged to derive age-scaled scores for five subscales: speech and auditory processing (SAP), environmental and auditory sensitivity (EAS), language/literacy/laterality (L/L/L), memory and attention (M&A), and pragmatic and social skills (PSS), each containing 6 to 9 distinct items. The subscales were further grouped into Language, Listening, Social, and Total aggregate (composite) scores, all of which were standardized with a population mean of 10 (SD = 3) based on British data (Barry et al. 2015). All ECLiPS subscales exhibit high test-retest reliability, with intraclass correlations (ICCs) above 0.8 (Barry and Moore 2021). Construct validity has been demonstrated through convergence with other established tests measuring similar skills, including the Children’s Auditory Processing Performance Scale (CHAPS; (Smoski et al. 1998)), the CCC-2 (Bishop 2006), and the Social Communication Questionnaire (Rutter et al. 2003). Although the ECLiPS provides scores for five latent factors related to listening (SAP, EAS, L/L/L, M&A, and PSS), these scores are not validated for diagnosing specific impairments in each area. Consequently, eligibility for inclusion in the LiD category was determined based on the ECLiPS Total composite score (below 7, which defines a cutoff of < 1 SD from the population mean). The Total composite score represents the overall degree of LiD (Barry and Moore 2021) and is appropriate for this purpose.

### Audiometry

Otoscopy was completed, and if necessary, cerumen was removed before audiometry. All audiometric tests were completed in a double-walled soundproof booth. Standard (0.25 - 8 kHz) and extended high-frequency (10-16 kHz) thresholds were measured using the manual Hughson-Westlake method for the range of 0.25 to 8 kHz at octave intervals and at four additional frequencies (10, 12.5, 14, and 16 kHz) using an Equinox audiometer (Interacoustics Inc., Middelfart, Denmark) with Sennheiser 300 HDA headphones. Participants with elevated thresholds (>20 dB HL) in either ear and at any standard clinical frequency were excluded and referred for further clinical care as appropriate.

### The Listening in Spatialized Noise-Sentences (LiSN-S) test

The Listening in Spatialized Noise – Sentences (LiSN-S) test (Cameron and Dillon 2007; Brown et al. 2010) measures the ability to listen and repeat simple, spoken sentences in the presence of distracting sentences, developed using the same child-friendly criteria as the BKB sentences (Bench et al. 1979). We selected LiSN-S as the auditory assessment tool for the SICLiD study because of its multifaceted approach to evaluating spatial and speech-hearing abilities, as well as its specific design for use in pediatric populations. The LiSN-S (US Edition (Brown et al. 2010)) was administered at each wave using a commercial CD played on a laptop, a task-specific soundcard, and Sennheiser HD 215 headphones. Participants were asked to repeat a series of target sentences, presented using virtual space generic head-related transfer functions (Humanski and Butler 1988), directly in front (0°: diotic) while ignoring two distracting speakers.

The LiSN-S has four listening conditions in which the distractors change voice (different or same as target) and/or position (0° and 90°) (Humanski and Butler 1988). The LiSN-S is adaptive; the level of the target speaker decreases or increases in SNR relative to the distracting speech if the listener responds correctly or incorrectly during up to 30 sentences in each condition. The speech reception threshold (SRT) represents 50% correct SNR for the condition. The Low Cue condition is where distractors are in the same voice as the target and in front (0°) relative to the listener. Distractors are in different voices and ±90° relative to the listener in the High Cue condition. Derived scores of the LiSN-S are the Talker Advantage and Spatial Advantage, so-called because each is the difference between SRTs from two conditions. This subtraction process should, to varying extent, separate auditory from cognitive influences (Cameron and Dillon 2007; Petley et al. 2021). Spatial Advantage thus represents the participant’s ability to separate the spatial source of the speech using purely auditory cues, while Talker Advantage demonstrates the participant’s ability to distinguish individual talkers, which involves both acoustic and linguistic information (Perrachione et al. 2011; Quinto et al. 2020). Test-retest comparisons for the four listening conditions of the LiSN-S showed significant improvement from the first test to the second but no differences in the advantage scores (Cameron et al. 2011).

A spatial “Pattern Score” is a quantitative clinical measure of the benefit of adding virtual spatial cues to the information in the LiSN-S (i.e., target and distracting stimuli presented diotically) (Cameron and Dillon 2011). The pattern score is calculated as follows.

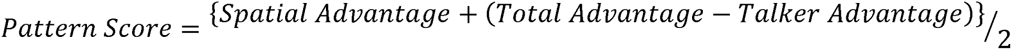

The Pattern Score has been used as both a diagnostic score and a treatment outcome for a specific auditory disorder termed Spatial Processing Disorder (SPD) (Cameron et al. 2011, 2012; Cameron et al. 2014). The age-specific cutoff value of the pattern score is determined as follows.

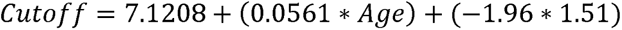

### Cognition (National Institutes of Health, NIH Toolbox)

Participants’ cognitive abilities were evaluated using the NIH Toolbox for the Assessment of Neurological and Behavioral Function, Cognition Domain (Weintraub et al. 2013a; Weintraub et al. 2013b). Participants completed testing either online or via an iPad app in a private sound-attenuated booth or a quiet room at each wave. The precise composition of the testing battery depended on participant age. Up to eight standardized cognitive instruments measuring different aspects of fluid or crystallized reasoning were utilized.

All participants in this study completed the Picture Vocabulary Test (PVT), Flanker Inhibitory Control and Attention Test (Flanker), Dimensional Change Card Sort test (DCCS), and Picture Sequence Memory test (PSMT). Each test produced a US age-corrected standardized score. The scores from all four tests were combined to calculate a single Early Childhood Composite (ECC), which provides a measure of overall cognitive skill for children aged six years and older. The PVT is an adaptive test that evaluates the participants’ vocabulary skills. Participants listen to an audio recording of a word and select one of four pictures that closely matches the word’s meaning. The Flanker assesses the participant’s ability to inhibit responses and focus their attention. Participants report the direction of a central visual stimulus in a series of five similar, flanking stimuli, where the flanking stimuli may either be congruent or incongruent. The DCCS tests cognitive flexibility (attention switching). Target and test card stimuli vary along two dimensions, shape, and color. Participants are asked to match test cards to the target card according to a specified dimension that varies for each trial. The PVT and DCCS scores both measure accuracy and reaction time. The PSMT assesses episodic memory by presenting illustrated objects and activities along with audio-recorded descriptive phrases. Participants are scored on the number of adjacent picture pairs correctly remembered over two learning trials. The length of the picture sequences varied from 6 to 18 pictures based on the participant’s age. The ECC score derived from these tests allows for a comprehensive evaluation of overall cognitive performance in early childhood.

Children at least 8 years of age qualify for additional subtests from the Toolbox, contributing to a Total Composite (TC) measure of general cognitive skill. These additional tests were administered to all children 8 years of age and older (i.e., all children for Waves 2 and 3). Fluid Composite measures include the list sorting working memory (LSWM), the pattern comparison processing speed (PCPS) tests, and the DCCS, Flanker, and PSMT. The LSWM subtest evaluates working memory by requiring participants to arrange visually and auditorily presented objects (food and animals) in order of size. In the PCPS subtest, participants were instructed to indicate as quickly as possible whether two visually presented cards were the same or different. Participants completed the Reading Recognition (RR) test in addition to the PVT for the Crystallized Composite measure. The RR requires participants to read words and letters aloud.

### Analysis

The analysis of the study was divided into three distinct parts, each corresponding to a specific aim of the research. In the first part, we evaluated the longitudinal changes in listening, auditory, and cognitive functions of children with LiD in comparison to TD children. In the second part, we developed a model to explain childhood listening difficulty using longitudinal data. Finally, the third part aimed to establish the long-term test-retest reliabilities of the NIH Cognition Toolbox, ECLiPS, and LiSN-S, using prospectively collected data.

In Part 1 of the analysis, we investigated differences between two groups, TD and LiD, on their demographics, auditory, and cognitive function. Demographic information included age, sex, race, ethnicity, and maternal education. Test scores of ECLiPS, LiSN-S, and NIH Cognition Toolbox were compared. We used mixed-effect models (Verbeke 1997; Fitzmaurice et al. 2012) to analyze the longitudinal data. These models can handle repeated measures and are flexible on missing data. We assumed the data were missing at random (MAR) and used Restricted Maximum Likelihood (REML) estimation. We tested the MAR assumption with Little’s test (Little 1988). A mixed-effect model has fixed effects and random effects. Fixed effects are variables that have a consistent effect on the outcome variable, while random effects are variables that have a varying effect on the outcome variable depending on the level of another variable. Mixed-effect models are useful for analyzing data that have repeated measurements or hierarchical structures. Mixed-effect models can use both dependent and independent variables of multiple time points by modeling the change in the outcome variable over time as a function of fixed and random effects. This allows accounting for within-subject correlation and between-subject variations in the data.

For each demographic characteristic, we created a mixed-effect model using group (TD vs. LiD), wave (Wave 1, 2, and 3), and interaction (Group x Wave) as fixed effects and participant as a random effect. Maternal education was collapsed into four groups: up to high school education (High school), some college education (College), Bachelor’s degree (Bachelor), and post-graduate education (Post-grad). Race was collapsed into two groups: white and non-white. Ethnicity was collapsed into two groups: Hispanic or Latino and others. Demographic characteristics that are statistically different between groups (p < .05) may confound the statistical analysis that compared behavioral testing results between TD and LiD. Therefore, such demographic variables were included in the analyses to control potential confounding.

Next, we evaluated longitudinal changes in listening and cognitive skills of TD and LiD children using the ECLiPS sub-scores and Total Scaled score, LiSN-S Advantage, Cue and Pattern scores, and NIH Cognition Toolbox sub-tests and composite scores. We constructed a mixed-effect model for each test, incorporating fixed effects for group (TD vs. LiD), wave (Wave 1 vs. 2), interaction (Group x Wave), and maternal education (a potential confounding variable), and participant as a random effect (Part 1a model). All analyses were conducted using standardized scores adjusted for age on the test date. We applied Benjamini and Hochberg (B-H) adjustment (Benjamini and Hochberg 1995) to the p-values for multiple comparisons, accounting for eight ECLiPS sub-scores, four LiSN-S scores, seven NIH Cognition Toolbox sub-scores, and four NIH Cognition Toolbox composite scores with a false discovery rate (FDR) of less than 5%. Wave 3 data were not included in these mixed-effect models due to the smaller sample size compared to other waves. To examine further longitudinal changes in Wave 3, we created separate mixed-effect models using data only from participants who completed all three waves (Part 1b model).

In Part 2 of the analysis, our objective was to identify the functional domains that contributed to LiD. We first tested whether a model developed in our prior cross-sectional study (Petley et al. 2021) could predict the ECLiPS Total Scaled score at Wave 2. The original model included the SCAN-3:C Composite Score, the LiSN-S Talker Advantage score, and the NIH PVT and DCCS scores as predictors of ECLiPS Total Scaled scores at Wave 1. Since the SCAN-3:C was not administered in Wave 2, this model included only the LiSN-S Talker Advantage score and NIH PVT and DCCS scores from Wave 2 as predictors. Next, we developed a new mixed-effect model using the longitudinal data to predict ECLiPS Total Scaled scores. This longitudinal model included both predictor variables and outcome variables from Waves 1 and 2 in a single mixed-effect model. Candidate predictors were maternal education, five LiSN-S scores (Cue, Advantage, and Pattern scores), and measures of cognitive function (either the seven NIH Cognition Toolbox subtests or the four NIH Toolbox composite scores), and the outcome variable was the ECLiPS Total Scaled score in both Waves 1 and 2. By modeling the trajectories of ECLiPS scores over time as a function of the predictors, this approach accounts for both within-subject correlations across waves and between-subject variations. The random effects of the model capture individual differences in trajectories, while the fixed effects estimate overall trends. Separate models were tested using the NIH Cognition Toolbox subtests or composite scores as candidate predictors to avoid including duplicate variables in the model. Owing to its smaller sample size, Wave 3 was omitted from this longitudinal model.

All candidate predictors were initially examined against the ECLiPS Total Scaled score through univariate analyses using a mixed-effect model, with participants used as a random effect and predictor variables and the interaction term (predictor x wave) as fixed effects. Only variables with p < 0.1 in the univariate analysis were included in the backward selection. Pairwise comparisons among the levels of significant variables were adjusted for using Tukey-Kramer multiple adjustment. Beta weights for each predictor variable were calculated to indicate the strength of their effects on the ECLiPS Total Scaled score.

The aim of Part 3 of the analysis was to evaluate the test-retest reliability of the measures. To achieve this, Intra-class correlation coefficients (ICC) and Pearson correlation coefficients were computed to investigate the consistency of the test outcomes between Waves 1 and 2 for all participants. Wave 3 data were excluded from the analysis due to the limited sample size. Furthermore, we calculated test-retest differences as another relevant metric. For each measure, we subtracted the Wave 2 score from the Wave 1 score for each participant. We then calculated the mean and standard deviation of these difference scores. To determine whether there were any variations in the correlation coefficients between the TD and LiD groups, the coefficients were calculated separately for each group, and z-tests were performed. To avoid type I errors, the p-values were adjusted for multiple comparisons using the B-H adjustment method (Benjamini and Hochberg 1995) with an FDR of less than 5%. B-H adjustment was separately applied to four LiSN-S scores, seven NIH Cognition Toolbox sub-scores, four NIH Cognition Toolbox composite scores, and eight ECLiPS sub-scores. All analyses used SAS version 9.4 (SAS Institute, Cary, N.C.), setting the two-sided significance level at 0.05.

## RESULTS

### LiD group associated with lower maternal education

Table 1 shows the number and demographics of participants in each group and wave. As Part 1 of our analysis, we compared the TD and LiD groups on their demographics, auditory, and cognitive function. Among the demographic variables, only maternal education differed significantly between the two groups, with a significant Group x Wave interaction observed (see Table 1, Supp. Table 1). Specifically, the TD group consistently had higher maternal education than the LiD group across all waves, and this difference increased from Wave 1 to Wave 3. To address the potential confounding effect of maternal education, we included it as a covariate in the subsequent analyses.

### Group differences in caregiver-reported listening skills were maintained at Wave 2

By design, the LiD group had lower ECLiPS scores than the TD group at Wave 1 (Figure 1A and Table 2) (Petley et al. 2021). This difference between the groups persisted at Wave 2 (Analysis Part 1a model). There was no significant change in ECLiPS scores across the two waves for either group. Furthermore, there was no significant Group x Wave interaction effect, indicating that the listening skills of both groups remained stable over time (Supp. Tables 2, 3).

**Figure 1.**
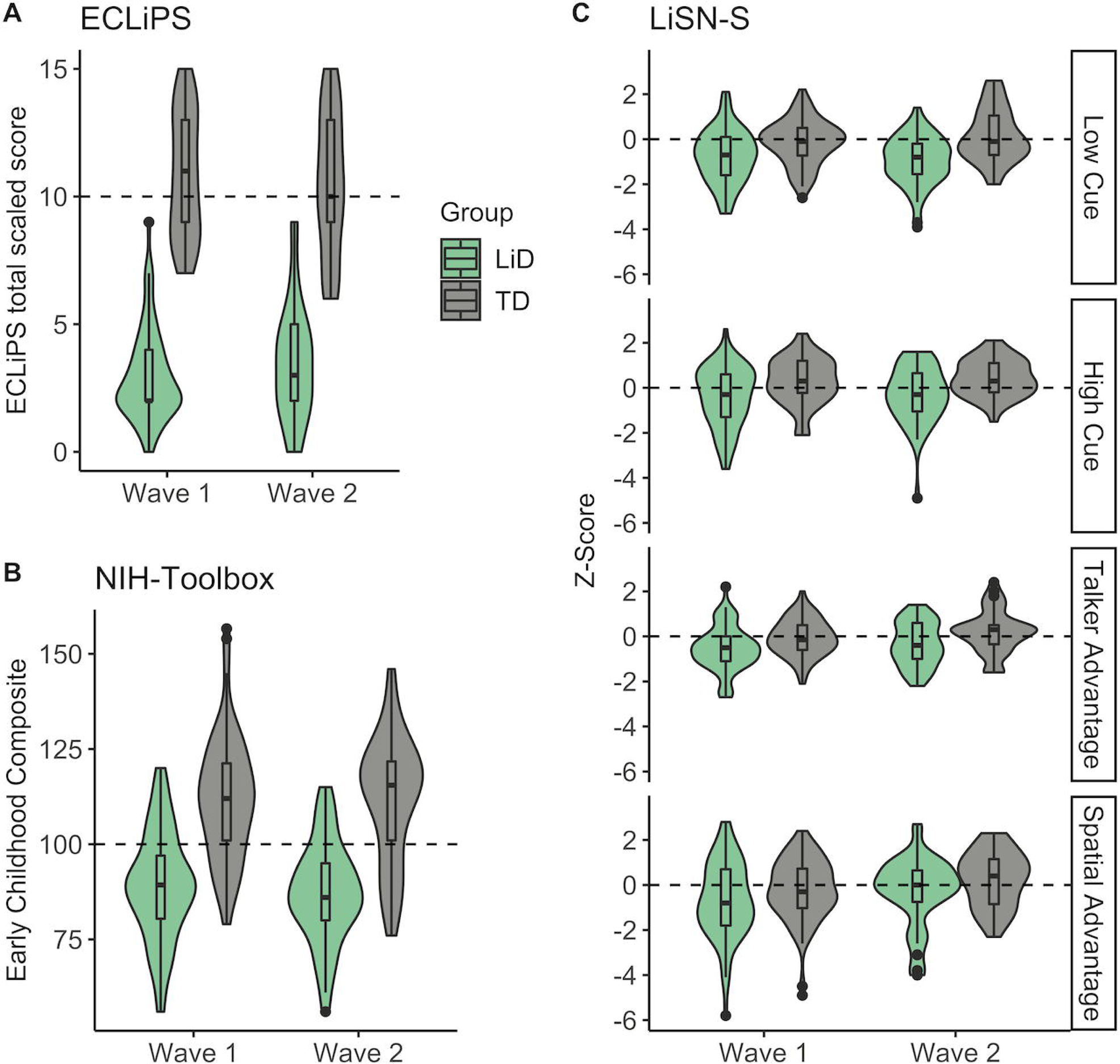
All test scores were consistently lower in the LiD group across Waves 1 and 2. **A.** Violin plots of ECLiPS Total Scaled scores showing the probability density of the data. Violin plots are overlaid with boxplots indicating each group’s median and interquartile range in Waves 1 and 2. The horizontal dashed line reflects the expected standard score (here, 10). Wave 3 data are not included in the figure due to the significantly smaller sample compared with Waves 1 and 2 (see Tables 1-3). **B.** Early Childhood Composite scores from the NIH Cognition Toolbox. **C.** LiSN-S Cue and Advantage scaled Z-Scores. LiD indicates listening difficulty; TD, typically developing.

**Table 2.**
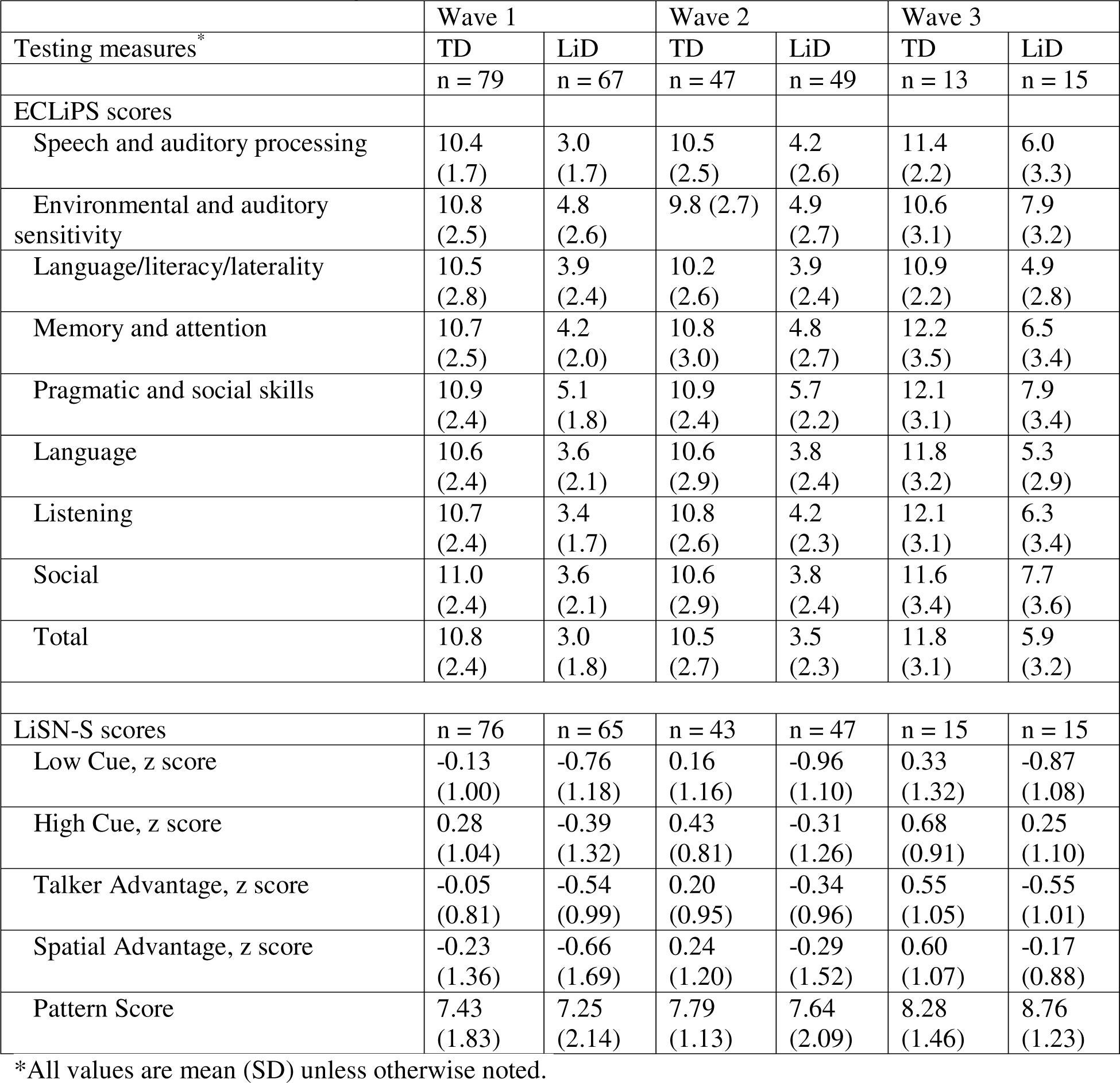
Auditory and listening test scores.

### LiD group had consistently lower cognitive scores

The LiD group scored significantly lower than the TD group on all seven sub-tests and four composite scores of the NIH Cognition Toolbox across Waves 1 and 2 (Figure 1B, Table 3, Supp. Tables 4, 5). Scores did not change significantly over time for either group as indicated by non-significant Wave or Group x Wave interaction effects.

**Table 3.** Cognitive test scores.

|  | Wave 1 |  | Wave 2 |  | Wave 3 |  |
| --- | --- | --- | --- | --- | --- | --- |
| Testing measures<sup>*</sup> | TD | LiD | TD | LiD | TD | LiD |
| NIH Cognition Toolbox Composite Scores | n = 56 | n = 49 | n = 42 | n = 45 | n = 15 | n = 15 |
| Picture vocabulary | 114 (14) | 98 (13) | 112 (14) | 93 (11) | 113 (15) | 91(16) |
| Flanker test | 101(15) | 90 (14) | 98 (12) | 86 (14) | 107 (15) | 80 (12) |
| List sorting working memory<sup>†</sup> | 110 (15) | 94 (14) | 110 (14) | 92 (13) | 117 (15) | 89 (12) |
| Dimensional change card sort test | 104 (15) | 89 (13) | 108 (19) | 90 (15) | 113 (21) | 89 (17) |
| Pattern comparison processing speed test<sup>†</sup> | 103 (22) | 87 (20) | 115 (22) | 86 (22) | 123 (19) | 90 (22) |
| Picture sequence memory test | 112 (19) | 95 (20) | 113 (18) | 99 (13) | 128 (17) | 95 (14) |
| Oral reading recognition test<sup>†</sup> | 107 (13) | 90 (12) | 110 (12) | 92 (11) | 106 (14) | 85 (11) |
| Fluid cognition composite<sup>†</sup> | 109 (18) | 85 (17) | 114 (17) | 84 (17) | 126 (14) | 81 (18) |
| Crystallized cognition composite<sup>†</sup> | 112 (14) | 93 (12) | 113 (13) | 91 (10) | 111 (15) | 86 (14) |
| Total composite<sup>†</sup> | 112 (15) | 87 (14) | 116 (15) | 85 (13) | 123 (15) | 80 (15) |
| Early Childhood composite | 112 (16) | 89 (14) | 112 (16) | 87 (13) | 122 (16) | 82 (18) |

<sup>\*</sup>All values are mean (SD) unless otherwise noted.
<sup>†</sup>Data missing for 9 children in Wave 1 (TD: n = 51, LiD: n = 45).

### LiD group had consistently lower skills in segregating sentences

The LiD group had lower LiSN-S z-scores than the TD group across waves (Figure 1C and Table 2, Supp. Table 5, 6). Spatial Advantage scores increased significantly from Wave 1 to Wave 2 for both groups. Other LiSN-S z-scores did not show any significant change over time. Spatial Pattern Scores were similar between groups and waves. Notably, fewer children met the diagnostic criteria for SPD based on Pattern Scores at Wave 2 than at Wave 1: only three children (3 LiD, 0 TD) compared to thirteen children at Wave 1 (7 LiD, 6 TD).

### Differences between the TD and LiD groups persisted through adolescence

We examined longitudinal changes in listening skills, speech-in-speech skills, and cognitive function across three waves based on 31 participants who completed all three waves (Figure 2). The TD group had higher ECLiPS sub-scores and Total Scaled scores than the LiD group (Analysis Part 1b model, Supp. Table 7), as in the Part 1a analysis of Waves 1 and 2 data. However, both groups had improved listening skills over time relative to the age-standardized scores. ECLiPS speech and auditory processing subscale score and composite Listening score were significantly higher in Wave 2 than Wave 1 across both groups. Moreover, ECLiPS Total Scaled scores in Wave 3 were significantly higher than Wave 1. In contrast to the listening skills, there was no significant change in cognitive function or speech-in-speech skills over time for either group. The TD group had significantly higher NIH Cognition Toolbox scores and two LiSN-S z scores (Low Cue and Talker Advantage) than the LiD group. Wave 1 participants who did not participate in Wave 2 dropped out randomly (Little’s p = 0.74). However, those who did not participate in Wave 3 were non-random (Little’s p = 0.01), suggesting systematic reasons for attrition. Maternal education was not included in the model due to the small sample size.

**Figure 2.**
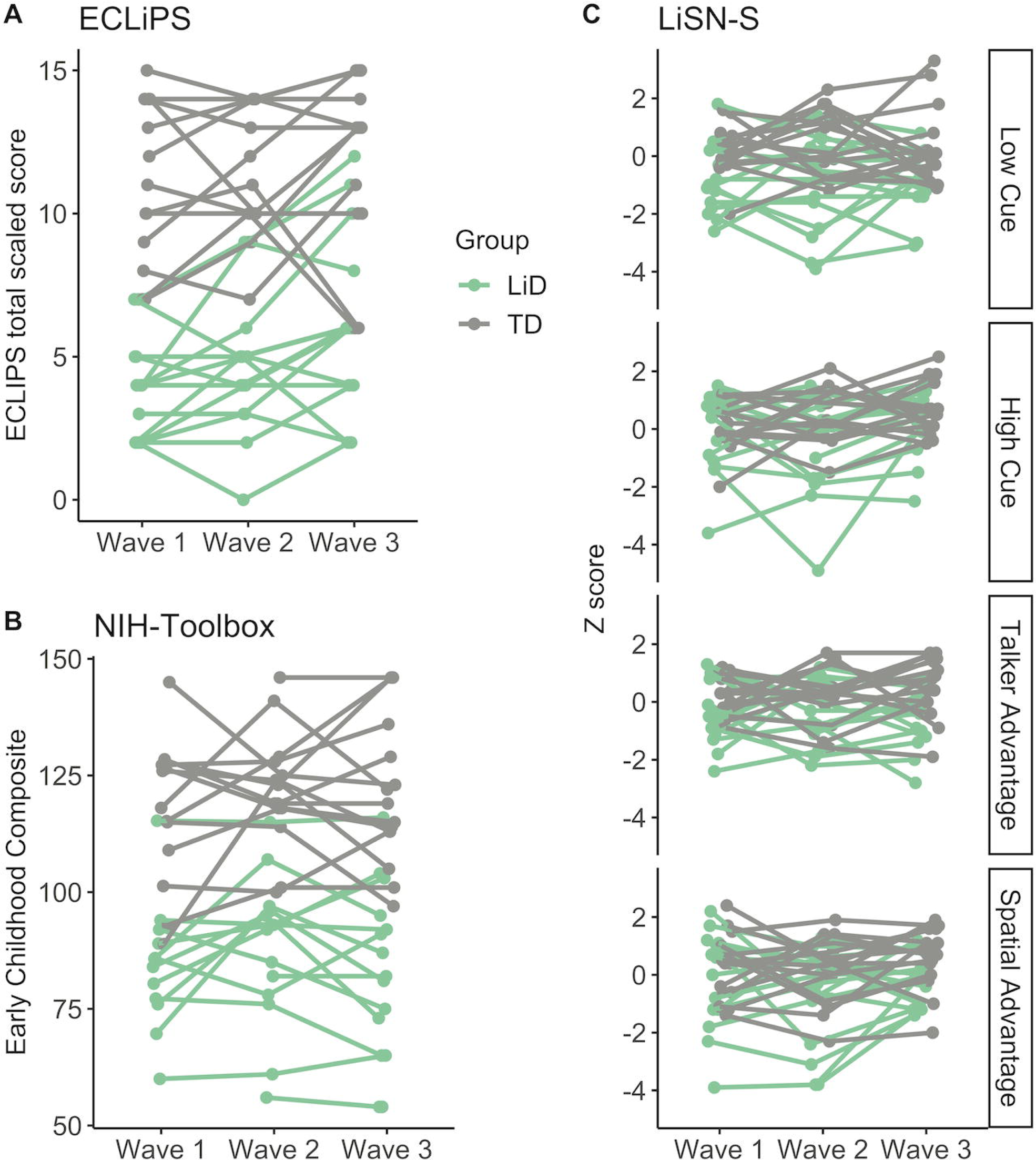
Individual data of Wave 3 participants showed improved listening skills over 3 waves. **A.** Line plots of ECLiPS total scaled scores across 4 years. **B.** Same as A for NIH-Toolbox Early Childhood Composite scaled scores. **C.** Same as A for LiSN-S Cue and Advantage scores. LiD indicates listening difficulty; TD, typically developing.

### Cognitive, sensory, and socioeconomic factors independently contributed to parent-reported listening skills

For Part 2 of the analysis, we compared two models to predict ECLiPS Total Scaled scores: one based on Wave 2 data only (Petley et al. 2021), and another based on longitudinal data from both Waves 1 and 2. The original model explained 42% of the variance of ECLiPS Total Scaled scores at Wave 2. Our new model explained 54.8% of the variance (χ²(2) = 48.21, p < 0.0001). The new model included three significant predictors: maternal education (β = .70, p = .0038), NIH total composite score (β = .49, p < .0001), and LiSN-S Spatial Advantage (β = .16, p = .0023). These results indicate that children’s listening skills during Waves 1 and 2 were independently influenced by their cognitive function, speech-in-speech skills, and maternal education level. Children whose mothers had a Bachelor’s degree scored higher on the ECLiPS than those whose mothers had a High school diploma by 3.1 points.

### ECLiPS, LiSN-S, and NIH Toolbox scores demonstrated reliability between Waves 1 and 2

As Part 3 of the analysis, we evaluated the test-retest reliability of the measures (Figure 3, Table 4, Supp. Table 8). The ECLiPS Total Scaled scores showed good reliability across Waves 1 and 2 with no differences between TD and LiD groups. The mean test-retest difference on the ECLiPS was 0.10, and the SD was 2.33. The NIH Toolbox sub-tests and composite scores also had significant and consistent correlations across waves (ICCs for each sub-test ranging from 0.48 to 0.82) and groups (p values from z-tests ranging from 0.07 to 0.92; Supp. Table 8). The mean test-retest differences ranged from −5.59 to 4.58, with the SDs ranging from 9.66 to 27.02 compared to the normalized mean score of 100. LiSN-S scores revealed a moderate but significant agreement between waves, with ICCs ranging from 0.28 to 0.42. The mean test-retest differences in standard deviation units were between 0.18 and 0.41, with SD between 1.26 and 2.13.

**Figure 3.**
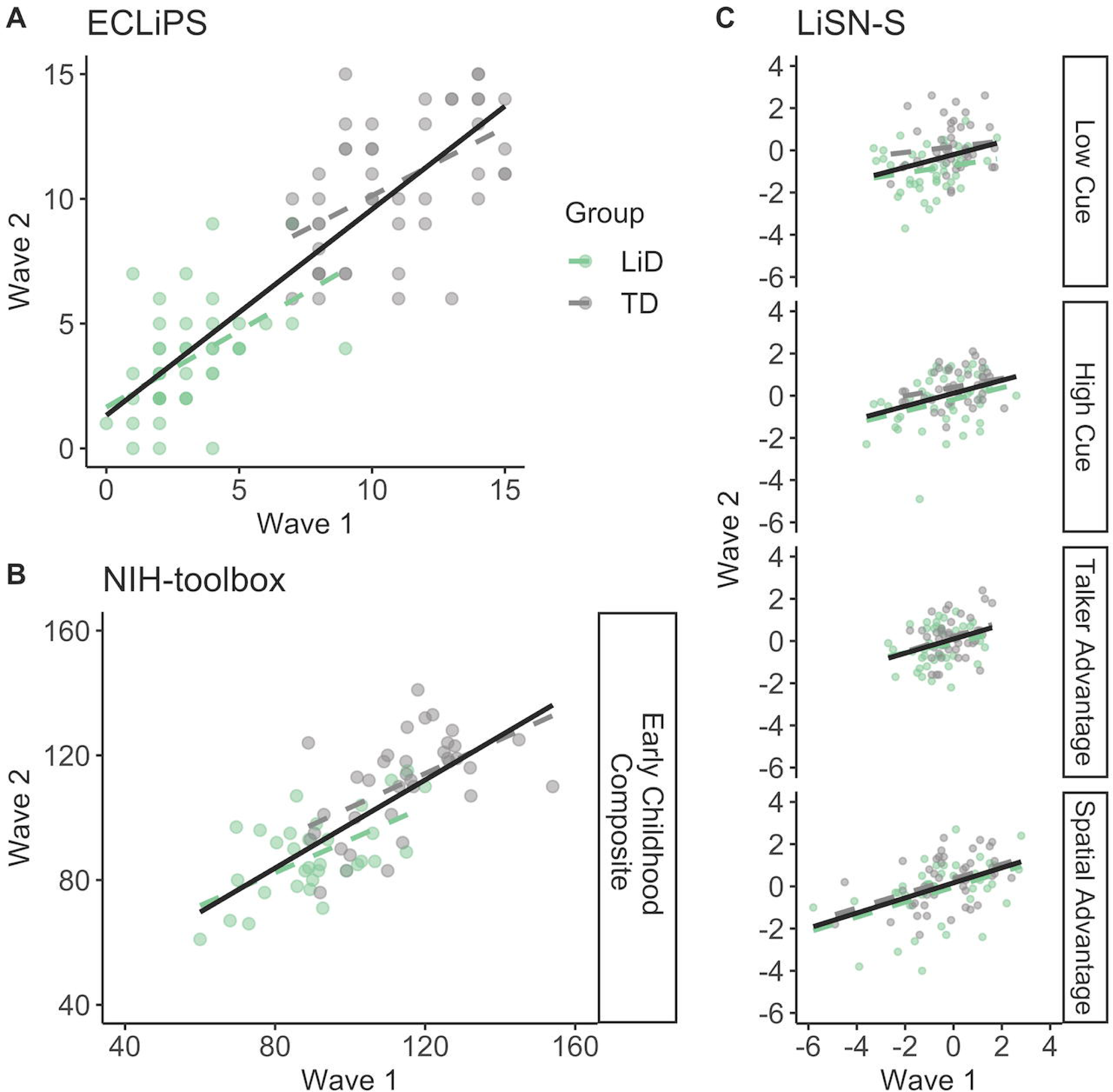
Listening skills and cognitive abilities showed good to excellent correlations between Waves 1 and 2. Auditory skills showed poor agreement between waves. **A.** Scatter plots of ECLiPS total scaled scores at Wave 1 and 2. Dashed lines show regression lines for each group. The black line indicates the regression line for the entire data combining groups. **B.** Same as A for Early Childhood Composite. **C.** Same as A for LiSN-S Cue and Advantage scores. LiD indicates listening difficulty; TD, typically developing.

**Table 4.** Pearson correlation coefficients, estimated reliability coefficients, and test-retest differences between waves 1 and 2.

|  | Pearson correlation coefficient | ICC (95% CI), Unadjusted | ICC (95% CI), Adjusted by Wave | Wave 2 – Wave 1 |  |  |  |
| --- | --- | --- | --- | --- | --- | --- | --- |
|  |  |  |  | N | Mean | SD | Mean/SD |
| ECLIPS total scaled score | 0.85 | 0.86 (0.80, 0.90) | 0.86 (0.80, 0.90) | 97 | 0.10 | 2.33 | 0.04 |
| NIH-toolbox age-corrected standard score |  |  |  |  |  |  |  |
| Picture vocabulary | 0.66 | 0.65 (0.51-0.77) | 0.65 (0.52-0.77) | 81 | -2.26 | 12.48 | -0.18 |
| Flanker test | 0.51 | 0.42 (0.26-0.60) | 0.48 (0.32-0.64) | 79 | -5.59 | 15.17 | -0.37 |
| List sorting working memory | 0.66 | 0.68 (0.53-0.79) | 0.68 (0.55-0.80) | 75 | -1.99 | 13.48 | -0.15 |
| Dimensional change card sort test | 0.48 | 0.53 (0.35-0.69) | 0.52 (0.35-0.69) | 79 | 0.12 | 17.97 | 0.01 |
| Pattern comparison processing speed test | 0.58 | 0.56 (0.40-0.71) | 0.56 (0.40-0.70) | 71 | 4.58 | 27.02 | 0.17 |
| Picture sequence memory test | 0.44 | 0.45 (0.27-0.64) | 0.45 (0.27-0.64) | 81 | 2.46 | 20.73 | 0.12 |
| Oral reading recognition test | 0.70 | 0.68 (0.54-0.79) | 0.70 (0.57-0.80) | 74 | 2.66 | 11.23 | 0.24 |
| Fluid cognition composite | 0.74 | 0.75 (0.64-0.84) | 0.76 (0.64-0.84) | 68 | -0.03 | 16.79 | 0.00 |
| Crystallized cognition composite | 0.80 | 0.79 (0.69-0.86) | 0.79 (0.69-0.86) | 72 | 0.84 | 9.66 | 0.09 |
| Total composite | 0.82 | 0.82 (0.73-0.88) | 0.82 (0.74-0.88) | 66 | 0.51 | 12.58 | 0.04 |
| Early childhood composite | 0.73 | 0.73 (0.61-0.82) | 0.74 (0.63-0.83) | 74 | -2.10 | 13.89 | -0.15 |
| LiSN-S, z score |  |  |  |  |  |  |  |
| Low Cue A | 0.29 | 0.27 (0.13-0.49) | 0.28 (0.13-0.49) | 89 | 0.41 | 2.13 | 0.19 |
| High Cue | 0.35 | 0.32 (0.17-0.52) | 0.33 (0.18-0.53) | 89 | 0.23 | 1.40 | 0.16 |
| Talker Advantage | 0.32 | 0.29 (0.14-0.51) | 0.33 (0.17-0.54) | 89 | 0.18 | 1.26 | 0.14 |
| Spatial Advantage | 0.45 | 0.39 (0.24-0.56) | 0.42 (0.27-0.58) | 89 | 0.33 | 1.69 | 0.20 |

## DISCUSSION

### Impact of study

We found that ECLiPS, LiSN-S, and NIH Toolbox scores were stable over time. Children with LiD, identified by the ECLiPS caregiver questionnaire, consistently performed more poorly than TD children on auditory and cognitive tasks across multiple years. Maternal education was a significant predictor of caregiver-reported listening skills, independent of auditory and cognitive skills. Socioeconomic factors may, therefore, play a crucial role in this complex condition. Aligning with our prior cross-sectional study, cognitive skills played a relatively larger role in the parent report of listening difficulty than auditory skills. The enduring nature of LiD has clinical implications for early detection and intervention for childhood LiD, which may have long-term effects on affected individuals.

### Socioeconomic factors in listening difficulties

This study is the first to show how socioeconomic factors affect reported listening skills in children with LiD after partially controlling for auditory and cognitive skills measured by the LiSN-S test and NIH Toolbox scores. Previous studies have linked maternal education to various aspects of child development, such as academic outcomes (Harding et al. 2015), language skills (Magnuson et al. 2009), attention problems (Hjern et al. 2010), and auditory brainstem responses (Skoe et al. 2013). It is therefore unsurprising that maternal education was associated with the degree of LiD in this study, given the known relation between LiD and these other developmental factors (Moore et al. 2018; Hunter et al. 2023). However, the finding is noteworthy because we controlled for cognitive and sensory functions using the LiSN-S test and NIH Toolbox scores, which suggests that maternal education may have an independent effect on LiD beyond these factors. Alternatively, maternal education may influence LiD through other factors. Future studies could further explore and parse the influence of socioeconomic factors. In the meantime, maternal education and socioeconomic factors may inform the risk of LiD and guide clinical management, including the frequency of follow-up and intervention. Clinicians could also provide support and guidance for parents with lower socioeconomic status who may face more challenges in accessing resources or implementing home-based activities for their children. They could also refer the children to other services, such as social workers or community programs, that can help them overcome disadvantages.

### Persistent listening challenges

This study provides evidence that children with LiD overall experience persistent listening challenges across time, as indicated by significantly lower mean ECLiPS scores compared to TD children across three waves of testing. Our analysis also accounts for the stability of scores for each participant over time, as we used mixed-effect models to evaluate the longitudinal score changes of each participant. The findings suggest that differences between TD and LiD groups are not due to a supra-normal performance of TD children, as their ECLiPS scores were consistent with other independent samples of children without LiD (Barry et al. 2015). The results are consistent with previous research (Del Zoppo et al. 2015) and underscore the need for early diagnosis and active management to minimize the long-term effects of LiD. Our findings also raise questions about the effectiveness of current management strategies for LiD, which may not adequately address the unique needs of these children. In reviews of medical records, we found that more than half the children with LiD had received specialist assessment and interventions before and/or during the study period. To better evaluate the efficacy of these interventions, future research should use quantitative outcome measures specific to each treatment. This will help identify the most effective strategies for addressing the persistent listening challenges experienced by children with LiD.

### Cognitive challenges

Cognitive challenges of LiD participants were persistent throughout the study period, as shown by our prior Wave 1 study (Petley et al. 2021; Stewart et al. 2022) and longitudinal data from this study. LiD participants performed more poorly than controls on all the NIH Toolbox subscale and composite scores. Modeling revealed that the NIH Total Composite score was the most predictive cognitive measure for reported LiD, surpassing any combination of NIH Toolbox subscales. Moreover, the NIH Total Composite score had a threefold higher beta weight than the next most predictive outcome, speech-in-speech repetition (LiSN-S Spatial Advantage). This finding underscores the substantial role of cognitive impairment in parent-reported LiD. It is worth noting that the LiD group in our study may have included a variety of underlying etiologies, as we used the ECLiPS Total Scaled score as an inclusion criterion. Consequently, our results are relevant to children with LiD as a whole, rather than to a specific underlying condition of LiD. To address distinct needs, individual children with specific deficits should receive evidence-based, targeted treatment (Dillon and Cameron 2021; Sharma et al. 2012; Musiek and Chermak 2007).

### Speech-in-speech listening

One purpose for using LiSN-S was to characterize the specific pattern of impairment in LiD participants using individual and derived scores of LiSN-S that represent different aspects of listening abilities. In this study (Part 1a), the TD group outperformed the LiD group on all age-adjusted LiSN-S scores. Our results thus highlight the significant challenges that LiD children face across multiple domains of listening ability. Interestingly, in the Part 1b analysis, children with LiD scored lower only in the Low Cue and Talker Advantage, similar to our previous findings in Wave 1 (Petley et al., 2021). Taken together, the data suggest that Low Cue and Talker Advantage may be more sensitive measures for differentiating TD and LiD than other LiSN-S scores for most children in this cohort.

Although the number of cases was small, the prevalence of SPD decreased at Wave 2, despite ongoing listening, auditory, and cognitive challenges in the LiD group and without any participants receiving specific treatment for SPD (Cameron et al. 2012). Furthermore, neither the Pattern score nor the frequency of SPD differed significantly between the TD and LiD groups, suggesting that, at least in this cohort, SPD does not represent the listening problems identified by the ECLiPS (Petley et al. 2021). Future research clarifying the clinical significance and natural history of SPD would be beneficial. Among LiSN-S scores, only the Spatial Advantage age-adjusted scores increased significantly from Waves 1 to 2 in both groups. This could be due to developmental changes or inconsistent normalization and is unlikely to be training effects after a 2-year separation between tests.

### Reliability

We believe our findings are based on reliable longitudinal data, as the current study utilized a large sample of children and had a 68% follow-up rate at Wave 2, much higher than a prior longitudinal study of childhood LiD with only a 13% follow-up response rate (Del Zoppo et al. 2015). Additionally, loss to follow-up at Wave 2 was random, suggesting a low likelihood of sampling bias. Findings from Waves 1 and 2 thus likely represent the typical progression of LiD. Unfortunately, due to COVID, the follow-up rate of Wave 3 dropped significantly, and sampling bias was introduced. However, analysis of Wave 3 participants showed overall longer-term results that were consistent with the primary analysis of Waves 1 and 2.

ECLiPS, LiSN-S, and NIH Toolbox scores all showed statistically significant correlations between Waves 1 and 2. These correlations were similar for the TD and LiD groups. The high reliability of the Toolbox scores observed in this study, even with long testing intervals of two years, seems to alleviate recent concerns regarding these scores (Taylor et al. 2022). This high reliability may be attributable to consistent testing procedures carried out by research staff at a single site, as opposed to testing across multiple sites (Taylor et al. 2022). Although LiSN-S age-adjusted scores were significantly correlated between Waves 1 and 2, the agreement between waves ranged from poor to good, and correlation coefficients were generally lower than those reported in a previous study (Cameron et al. 2011). The higher retest reliability for that previous study may be explained by shorter testing intervals (2-4 months) and older (above 12 years old) participants (Cameron et al. 2011).

### Limitation

Due to the small sample size of Wave 3, we ran a separate analysis focusing on longitudinal changes of Wave 3 participants to reduce sampling bias. Though this method reduced bias, the analysis remained underpowered. Despite this limitation, our overall research findings are significant because we showed long-lasting challenges of childhood LiD that may influence clinical management strategy, and we identified a social factor, maternal education, that independently contributes to reported LiD.

### Conclusion

Children with LiD and clinically normal audiograms have persistent auditory, listening, and cognitive challenges through at least early adolescence. The degree of LiD can be independently predicted by maternal education, cognitive processing, and spatial listening skills with a relatively larger influence of cognitive processing over other factors.

## Data sharing statement

The data that support the findings of this study are available from the corresponding author upon reasonable request.

## ACKNOWLEDGEMENTS

This research was supported by grant 5R01DC014078 from the National Institute of Deafness and other Communication Disorders (DRM), 2UL1TR001425-05A1 by the National Center for Advancing Translational Sciences of the National Institutes of Health (KK), and by the Cincinnati Children’s Research Foundation. DRM received support from NIHR Manchester Biomedical Research Centre. We would like to acknowledge that an earlier version of this manuscript was published as a preprint on medRxiv (https://doi.org/10.1101/2022.08.11.22278673).

K.K. analyzed and interpreted data and cowrote the article. L.L. analyzed and interpreted data and was involved in critical revisions. L.P. was involved in critical revisions. N.C. and A.P. collected data. M.B. analyzed and interpreted data and was involved in critical revisions. C.M.B., L.M.Z, and L.L.H. collected data and were involved in critical revisions. D.R.M. designed the study, analyzed and interpreted data, and cowrote the article.

