## Supplemental Tables 1-8 for "Childhood listening and associated cognitive difficulties persist into adolescence"

Supplemental Table 1. The p-values from mixed effect models for demographics

|  | Wave | Group | Group* Wave |
| --- | --- | --- | --- |
| Age at wave* | <.0001 | 0.3724 | 0.0074 |
| Sex^ | 1.0000 | 0.1538 | 1.0000 |
| Maternal education^ | 0.8763 | <.0001 | <.0001 |
| Race^ | 1.0000 | 0.1656 | 1.0000 |
| Ethnicity^ | 1.0000 | 0.6770 | 1.0000 |

* Linear mixed model; ^Generalized mixed model

Supplemental Table 2. The p-values from mixed effect models for ECLiPS scores (Wave, Group, maternal education, and Wave*Group)

|  | The mixed model with wave 1 & 2 | | | |
| --- | --- | --- | --- | --- |
|  | Wave | Group | Wave*Group | Maternal education |
| ECLiPS Total Scaled score | 0.6254 | <.0001 | 0.1389 | 0.3652 |
| ECLiPS subscores |  |  |  |  |
| SAP scaled score | 0.0096 | <.0001 | 0.0274 | 0.9530 |
| EAS scaled score | 0.0140 | <.0001 | 0.0205 | 0.3131 |
| LLL | 0.5788 | <.0001 | 0.7481 | 0.4016 |
| PSS | 0.1361 | <.0001 | 0.2477 | 0.7586 |
| MA | 0.2491 | <.0001 | 0.4962 | 0.4493 |
| Listening | 0.0560 | <.0001 | 0.2324 | 0.8131 |
| Language | 0.6628 | <.0001 | 0.8943 | 0.4279 |
| Social | 0.3385 | <.0001 | 0.0372 | 0.3766 |

Supplemental Table 3. Benjamini and Hochberg (B-H) adjusted critical values for ECLiPS subscores

|  | 8 ECLiPS subscores |
| --- | --- |
| Rank | B-H critical value, (i/m)Q |
| 1 | 0.00625 |
| 2 | 0.0125 |
| 3 | 0.01875 |
| 4 | 0.025 |
| 5 | 0.03125 |
| 6 | 0.0375 |
| 7 | 0.04375 |
| 8 | 0.05 |

i = the individual p-value’s rank; m = total number of tests; Q = the false discovery rate 0.05

Supplemental Table 4. The p-values from mixed effect models for NIH scores (Wave, Group, maternal education, and Wave*Group)

|  | Wave 1& 2 | | | |
| --- | --- | --- | --- | --- |
|  | Wave | Group | Group* Wave | Maternal education |
| NIH-toolbox age-corrected standard score |  |  |  |  |
| Picture vocabulary | 0.0456 | <.0001 | 0.4236 | 0.1238 |
| Flanker test | 0.0113 | 0.0009 | 0.7715 | 0.0005 |
| List sorting working memory | 0.3141 | <.0001 | 0.6117 | 0.0089 |
| Dimensional change card sort test | 0.4980 | <.0001 | 0.5117 | 0.3680 |
| Pattern comparison processing speed test | 0.1459 | <.0001 | 0.0113 | 0.3852 |
| Picture sequence memory test | 0.3241 | <.0001 | 0.3179 | 0.0006 |
| Oral reading recognition test | 0.0557 | <.0001 | 0.7157 | 0.0067 |
| Fluid cognition composite | 0.9147 | <.0001 | 0.1419 | 0.0004 |
| Crystallized cognition composite | 0.7008 | <.0001 | 0.6402 | 0.0142 |
| Total composite | 0.8833 | <.0001 | 0.1325 | <.0001 |
| Early childhood composite | 0.2897 | <.0001 | 0.9545 | 0.0002 |

Supplemental Table 5. B-H adjusted critical values for NIH-TB sub scores and LiSN-S scores

|  | 7 NIH-TB sub-scores | 4 LiSN-S scores |
| --- | --- | --- |
| Rank | B-H critical value, (i/m)Q | B-H critical value, (i/m)Q |
| 1 | 0.0071 | 0.0125 |
| 2 | 0.0143 | 0.025 |
| 3 | 0.0214 | 0.0375 |
| 4 | 0.0286 | 0.05 |
| 5 | 0.0357 |  |
| 6 | 0.0429 |  |
| 7 | 0.05 |  |

i = the individual p-value’s rank; m = total number of tests; Q = the false discovery rate 0.05

Supplemental Table 6. The p-values from mixed effect models for LiSN-S scores and Pattern score (Wave, Group, maternal education, and Wave*Group)

|  | Wave 1& 2 | | | |
| --- | --- | --- | --- | --- |
|  | Wave | Group | Group* Wave | Maternal education |
| LiSN-S, z score |  |  |  |  |
| Low cue | 0.6099 | <.0001 | 0.1493 | 0.0709 |
| High cue | 0.2406 | 0.0014 | 0.8433 | 0.3021 |
| Talker advantage | 0.0236 | 0.0121 | 0.9183 | 0.0618 |
| Spatial advantage | <.0001 | 0.0332 | 0.7206 | 0.1392 |
| Pattern score | 0.1057 | 0.4140 | 0.8365 | 0.5719 |

Supplemental Table 7. The p-values from mixed effect models for 3 waves (Wave, Group, and Wave*Group)

|  | The mixed model with three waves | | |
| --- | --- | --- | --- |
|  | Wave | Group | Wave*Group |
| ECLIPS total scaled score | 0.0275 | <.0001 | 0.7072 |
| SAP scaled score | 0.0043 | <.0001 | 0.1750 |
| EAS scaled score | 0.4351 | <.0001 | 0.2439 |
| LLL | 0.4636 | <.0001 | 0.6152 |
| PSS | 0.0465 | <.0001 | 0.8189 |
| MA | 0.0254 | <.0001 | 0.4905 |
| Listening | 0.0030 | <.0001 | 0.9319 |
| Language | 0.0521 | <.0001 | 0.1991 |
| Social | 0.9581 | <.0001 | 0.3756 |
| LiSN-S, z score |  |  |  |
| Low cue | 0.9388 | 0.0007 | 0.7279 |
| High cue | 0.0531 | 0.0914 | 0.4615 |
| Talker advantage | 0.8151 | 0.0026 | 0.4454 |
| Spatial advantage | 0.0972 | 0.0427 | 0.9104 |
| NIH-toolbox age-corrected standard score |  |  |  |
| Picture vocabulary | 0.4106 | <.0001 | 0.2161 |
| Flanker test | 0.3553 | <.0001 | 0.1680 |
| List sorting working memory | 0.7771 | <.0001 | 0.3232 |
| Dimensional change card sort test | 0.6842 | <.0001 | 0.4339 |
| Pattern comparison processing speed test | 0.0585 | 0.0002 | 0.0453 |
| Picture sequence memory test | 0.0174 | <.0001 | 0.583 |
| Oral reading recognition test | 0.0389 | <.0001 | 0.3950 |
| Fluid cognition composite | 0.1327 | <.0001 | 0.1236 |
| Crystallized cognition composite | 0.0181 | <.0001 | 0.2048 |
| Total composite | 0.0311 | <.0001 | 0.5832 |
| Early childhood composite | 0.3687 | <.0001 | 0.7537 |

Supplemental Table 8. Outcome Pearson correlation coefficients (r) between wave 1 and wave 2

|  | All | TD | LiD | r_1_-r_2_ | z-test, p-value |
| --- | --- | --- | --- | --- | --- |
| ECLIPS total scaled score | 0.85, p <.0001 | 0.52, p= 0.0002, n=47 | 0.52, p= 0.0002, n=46 | 0 | z=0, p=1.000 |
| LiSN-S, z score |  |  |  |  |  |
| Low cue | 0.29, p= 0.0068 | 0.12, p= 0.4588, n=43 | 0.20, p= 0.1954, n=42 | -0.08 | z=0.3651, p=0.7151 |
| High cue | 0.35, p= 0.0009 | 0.27, p= 0.0820, n=43 | 0.32, p= 0.0412, n=42 | -0.05 | z=02434, p=0.8077 |
| Talker advantage | 0.32, p= 0.0029 | 0.30, p= 0.0516, n=43 | 0.25, p= 0.1155, n=42 | 0.05 | z=0.2404, p=0.8100 |
| Spatial advantage | 0.45, p <.0001 | 0.45, p= 0.0027, n=43 | 0.44, p= 0.0034, n=42 | 0.01 | z=0.05541, p=0.9558 |
| LiSN-S, raw score |  |  |  |  |  |
| Low cue | 0.33, p= 0.0024 | 0.29, p= 0.0583, n=43 | 0.18, p= 0.2654, n=42 | 0.11 | z=0.5181, p=0.6044 |
| High cue | 0.46, p <.0001 | 0.48, p= 0.0012, n=43 | 0.42, p= 0.0056, n=42 | 0.06 | z=0.3346, p=0.7379 |
| Talker advantage | 0.38, p= 0.0004 | 0.37, p= 0.0150, n=43 | 0.32, p= 0.0360, n=42 | 0.05 | z=0.2523, p=0.8008 |
| Spatial advantage | 0.44, p <.0001 | 0.48, p= 0.0012, n=43 | 0.44, p= 0.0039, n=42 | 0.04 | z=0.2255, p=0.8216 |
| Pattern score | 0.21, p =0.0492 | 0.18, p= 0.2274, n=46 | 0.24, p= 0.1138, n=43 | -0.05 | z=0.3001, p=0.7642 |
| NIH-toolbox age-corrected standard score |  |  |  |  |  |
| Picture vocabulary | 0.66, p <.0001 | 0.58, p= 0.0002, n=36 | 0.42, p= 0.0163, n=32 | 0.16 | z=0.8438, p=0.3988 |
| Flanker test | 0.51, p <.0001 | 0.35, p= 0.0356, n=36 | 0.51, p= 0.0027, n=32 | -0.16 | z=0.7751, p=0.4383 |
| List sorting working memory | 0.66, p <.0001 | 0.50, p= 0.0039, n=31 | 0.52, p= 0.0033, n=30 | -0.02 | z=0.1002, p=0.9202 |
| Dimensional change card sort test | 0.48, p <.0001 | 0.35, p= 0.0380, n=36 | 0.30, p= 0.0905, n=32 | 0.05 | z=0.2197, p=0.8261 |
| Pattern comparison processing speed test | 0.58, p <.0001 | 0.59, p= 0.0005, n=31 | 0.32, p= 0.0832, n=30 | 0.27 | z=1.2829, p=0.1995 |
| Picture sequence memory test | 0.44, p= 0.0002 | 0.17, p= 0.3288, n=36 | 0.56, p= 0.0008, n=32 | -0.39 | z=1.8118, p=0.0700 |
| Oral reading recognition test | 0.70, p <.0001 | 0.54, p= 0.0019, n=31 | 0.61, p= 0.0004, n=30 | -0.07 | z=0.3884, p=0.6977 |
| Fluid cognition composite | 0.74, p <.0001 | 0.61, p= 0.0002, n=31 | 0.55, p= 0.0015, n=30 | 0.06 | z=0.3357, p=0.7371 |
| Crystallized cognition composite | 0.80, p <.0001 | 0.74, p <.0001, n=31 | 0.56, p= 0.0014, n=30 | 0.18 | z=1.1777, p=0.2389 |
| Total composite | 0.82, p <.0001 | 0.72, p<.0001, n=31 | 0.58, p= 0.0007, n=30 | 0.14 | z=0.9090, p=0.3633 |
| Early childhood composite | 0.73, p <.0001 | 0.54, p= 0.0008, n=36 | 0.59, p= 0.0003, n=32 | -0.05 | z=0.2888, p=0.7727 |
